# GPT-4 and Neurologists in Screening for Mild Cognitive Impairment in the Elderly: A Comparative Analysis Study

**DOI:** 10.1101/2023.12.02.23299327

**Authors:** Hao Yang, Ruihan Wang, Changyu Wang, Hui Gao, Hanlin Cai, Fengying Zhang, Jialin Liu, Siru Liu

## Abstract

This study evaluates the efficacy of GPT-4 in screening for Mild Cognitive Impairment (MCI) in the elderly, comparing it with junior neurologists. MCI is a precursor to dementia, presenting a significant public health concern due to the rising global aging population. With over 55 million people affected by dementia worldwide, early detection is essential for timely intervention. Common screening tools, while effective, are resource-intensive, highlighting the need for more efficient methods. The study used an exploratory design with 174 participants, comparing the performance of GPT-4 against three junior neurologists. The GPT-4 model was trained using a set of language analysis indicators to evaluate the severity of MCI. Participants’ test texts and voices were grouped and independently assessed by the neurologists and the GPT-4 model. The neurologists and the GPT-4 model independently assessed the participants’ test corpus. The neurologists assessed both the text and voice of the test, while the GPT model assessed the text only. Results showed that the GPT-4 model had higher accuracy (0.81) compared to the neurologists (ranging from 0.41 to 0.49). GPT-4 demonstrated better discrimination of MCI with significant statistical difference (p < 0.001). The study also developed a clinical risk assessment nomogram based on the top ten weighted features from GPT-4’s analysis, aiding in MCI patient evaluation. In conclusion, the GPT-4 model shows promise as a diagnostic aid for MCI, potentially improving patient outcomes and reducing healthcare burdens. However, its practical applicability in real-world scenarios requires further investigation and clinical validation.

## Introduction

The growing global aging population has brought Mild Cognitive Impairment (MCI) into sharp focus as a critical public health issue. MCI is increasingly recognized as the precursor stage to dementia, with a considerable risk of progression to advanced dementia [1]. Unfortunately, the absence of specific pharmaceutical treatments for MCI underscores the critical need for early detection and timely intervention [2,3]. The World Alzheimer Report 2023 emphasizes the growing challenge of dementia, with over 55 million people affected worldwide and a rising incidence rate [4,5]. The World Health Organization (WHO) estimates that by 2030, the number of people with dementia will reach 75 million, with the associated care costs expected to soar to 2 trillion USD. This escalation poses substantial societal and economic burdens [6]. Thus, early detection of cognitive impairments is vital for providing appropriate interventions and care, particularly for the aging population [7].

Early and accurate identification of cognitive changes using straightforward tools is key to guiding individuals towards more comprehensive neurocognitive evaluations and the formulation of treatment plans. Common screening instruments include the Hasegawa Dementia Scale-revised (HDS-R) [8,9], the Mini-Mental State Examination (MMSE) [8,10], Addenbrooke’s Cognitive Examination (ACE)-revised [8,11], and the Montreal Cognitive Assessment (MoCA) [12]. While these assessments have been adapted for simplicity, they still impose significant demands on healthcare providers and financial resources. Challenges such as healthcare provider shortages and time and financial constraints complicate these assessments. Consequently, there is a pressing need for an intelligent dementia screening tool that can alleviate the strain on healthcare systems and enable early and effective management of MCI [13].

Recent advancements in AI offer promise in addressing these challenges. AI’s role in healthcare is expanding, particularly in diagnostics and decision-making [14]. Current early screening approaches for cognitive impairments include using digital tools like tablet computers [11,12,15], virtual reality [16–18], and machine interactions with robots. These methods contrast with the more complex procedures typically conducted by physicians [19]. AI algorithms analyze vast patient data, enhancing the accuracy of clinical decisions and improving health outcomes. This technology also plays a crucial role in improving patient safety, optimizing health outcomes, and transforming clinical decision-making [20]. AI-based automatic screening for MCI offers the promise of enhancing diagnostic accuracy while reducing healthcare costs. Detecting pathological changes at their earliest stages could improve the outcomes of both pharmacological and non-pharmacological treatments. Therefore, developing a more sensitive, less invasive, cost-effective, and user-friendly diagnostic tool for MCI is of utmost importance.

This study aims to explore the potential of AI, particularly GPT models like ChatGPT, in the early detection of MCI. Given AI’s burgeoning role in healthcare, this research seeks to evaluate the accuracy, reliability, and practicality of using these models for MCI screening. This approach could provide an efficient and accessible method for early MCI detection, offering significant implications for healthcare systems and patient outcomes globally.

## Methods

### Study Design

This research was an exploratory study involving patients with MCI and normal cognition (CN) older people.

### Dataset Descriptions

We included a total of 174 subjects from the DementiaBank English Protocol Delaware Corpus [16] and the DementiaBank English Pitt Corpus [17,18]. 66 MCI subjects and 108 CN subjects were included [21].

### GPT4 Model

We employed the pre-existing GPT training model. The training process can be outlined as follows: Drawing on information from previously published studies, we meticulously defined essential language analysis indicators, encompassing lexical features, syntactic and grammatical attributes, and semantic characteristics, resulting in a set of seven core indicators (refer to Supplementary Materials). Upon completion of Step 1 training, participants in the study received Feedback 1, comprising more comprehensive features related to Mild Cognitive Impairment (MCI). These insights from the feedback were thoughtfully considered in new crafting prompts for Step 2. Step 2 was devised based on feedback data acquired from Feedback 2, which was utilized to generate entirely new prompts. In this phase, we extracted 21 indicators for evaluating the severity of MCI from MCI-related information (see Supplementary Materials) and established a scoring system to effectively measure the severity. Subsequently, GPT-4 gained the capability to autonomously design prompts, resulting in the creation of three distinct prompts that were later amalgamated. Moreover, the 21 indicators obtained from GPT-4 underwent statistical analysis. This ultimately culminated in the development of the GPT-4 Model [21].

### Grouping of Test Materials

A cleaned 174 copies of the test text and voice material were put together, of which 66 were mildly cognitively impaired and 108 were cognitively normal participants. These participants were included in each sample group so that neurologists could assess their cognitive status. The test texts and voice in each group were randomly numbered, and there were 4 groups. Each psychiatrist independently evaluated the 4 groups of test text material, and each group was evaluated approximately 7 days apart to minimize subjective bias on the part of the evaluator.

### Neurologist Selection

In this study, a purposive sampling method was used to select neurologists to ensure an accurate representation of the target population. As screening for cognitive impairment is typically carried out by junior neurologists and neurologist assistants, specific criteria were used to select participants. These criteria included the following qualifications 1. less than 5 years of professional experience in the field; 2. possession of an MD degree in neurology; 3. affiliation with A-level tertiary hospitals, which represent the highest-ranking healthcare institutions in China; 4. proficiency in English; and 5. willingness to participate in the study. Ultimately, three neurologists met these criteria and were selected. All of the selected neurologists have the necessary skills to detect mild cognitive impairment and joined the study group after qualifying.

### Statistics Analysis

Descriptive statistics were conducted to analyze each variable in each group. The calculations included mean, standard deviation, median, interquartile range, minimum and maximum values. Normal distribution and homogeneity of variance tests were also performed for all variables. For the analysis of continuous variables, one-way analysis of variance (ANOVA) [22] was used as the method of statistical analysis between groups. Chi-squared tests were used for statistical analysis to compare classification results between different neurologists and the GPT-4. All statistical calculations were performed using the Python programming language, implementing the SciPy and NumPy libraries to perform one-way ANOVA and chi-squared tests. Finally, violin plots were used to visually compare the level of concentration and dispersion for each variable.

### Ethical

The data utilized in this research was acquired from the publicly available DementiaBank dataset archived by TalkBank. TalkBank adheres to its own Code of Ethics, which supplements recognized professional guidelines such as the American Psychological Association Code of Ethics and the American Anthropological Association Code of Ethics [23], without replacing them. Importantly, the data does not include personal patient information and thus does not necessitate ethical approval or individual patient consent. Furthermore, all protected health information was appropriately anonymized to comply with data protection regulations.

## Results

A total of three junior neurologists, one male and two females, participated in the study. Their mean age was 27 years (±2.8) and their mean clinical experience in neurology was 12 months. The characteristics of the neurologists are shown in Table 1.

**Table 1.** Characteristic of neurologists’ participants in the study.

| Characteristic | Neurologists |
| --- | --- |
| Gender |  |
| Male | 1 |
| Female | 2 |
| Age (years) | 25, 25, 31 (27±2.8) |
| Degree | M Med, MD, MD Candidate |
| Years of clinical experience (months) | 2, 6, 24 (10.7±7.4) |

### Model performance

We used the pre-trained completed GPT4 model and Table 2 shows the results of the GPT4 model. Its F1 scores for the test and training sets are 0.88 and 0.77, with accuracies of 0.92 and 0.81, respectively. The receiver operating characteristic curve is 0.86, reflecting the model’s ability to discriminate between the data on the test set. The DCA curve [24] for the GPT-4 model shows a significant net benefit, suggesting that the use of the model in medical decision making can have practical benefits. In particular, the GPT-4 model demonstrated excellent performance over a specific range of decision thresholds, providing a significant advantage over no-action or other potential models.

**Table 2.** The classification results of the GPT-4 model on the training and test sets.

| Dataset | TP | FN | FP | TN | SEN | SPE | F1-Score | Accuracy |
| --- | --- | --- | --- | --- | --- | --- | --- | --- |
| Training set | 38 | 6 | 4 | 74 | 0.86 | 0.95 | 0.88 | 0.92 |
| Test set | 17 | 5 | 5 | 25 | 0.77 | 0.83 | 0.77 | 0.81 |

### Top 10 feature for determining MCI

Based on the features returned by the GPT-4 results, we built a logistic model to obtain the feature weights. This represents the contribution of the different features in the model to the discrimination of MCI. These coef_ values were then sorted to determine which features contributed most to the discrimination of MCI. Finally, the top ten features with the most significant contributions are shown in Figure 3. These features have significant weights, highlighting their central role in the predictive ability of the GPT-4 model for the discrimination of MCI.

**Figure 1.**
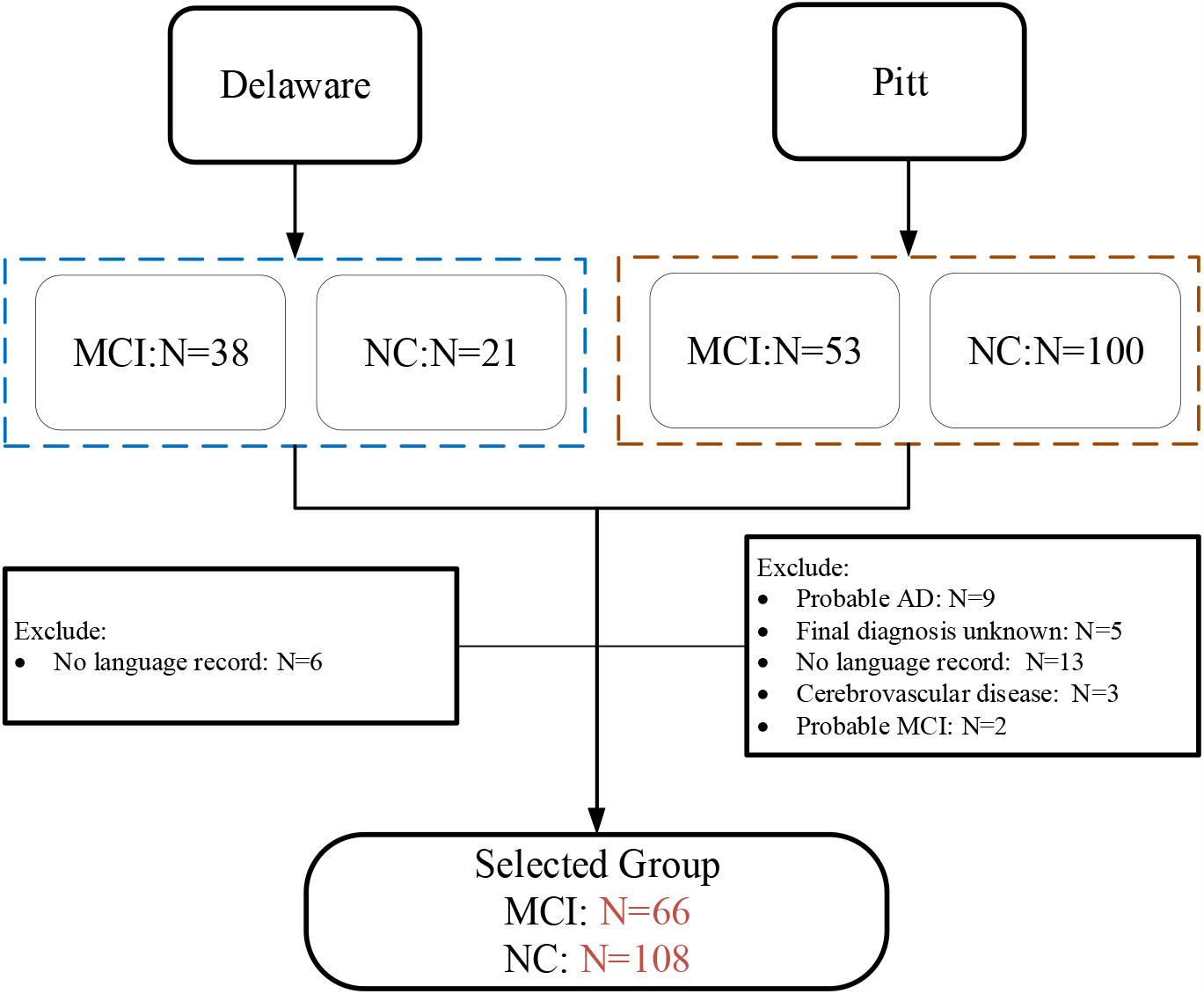
Participant inclusion and exclusion

**Figure 2.**
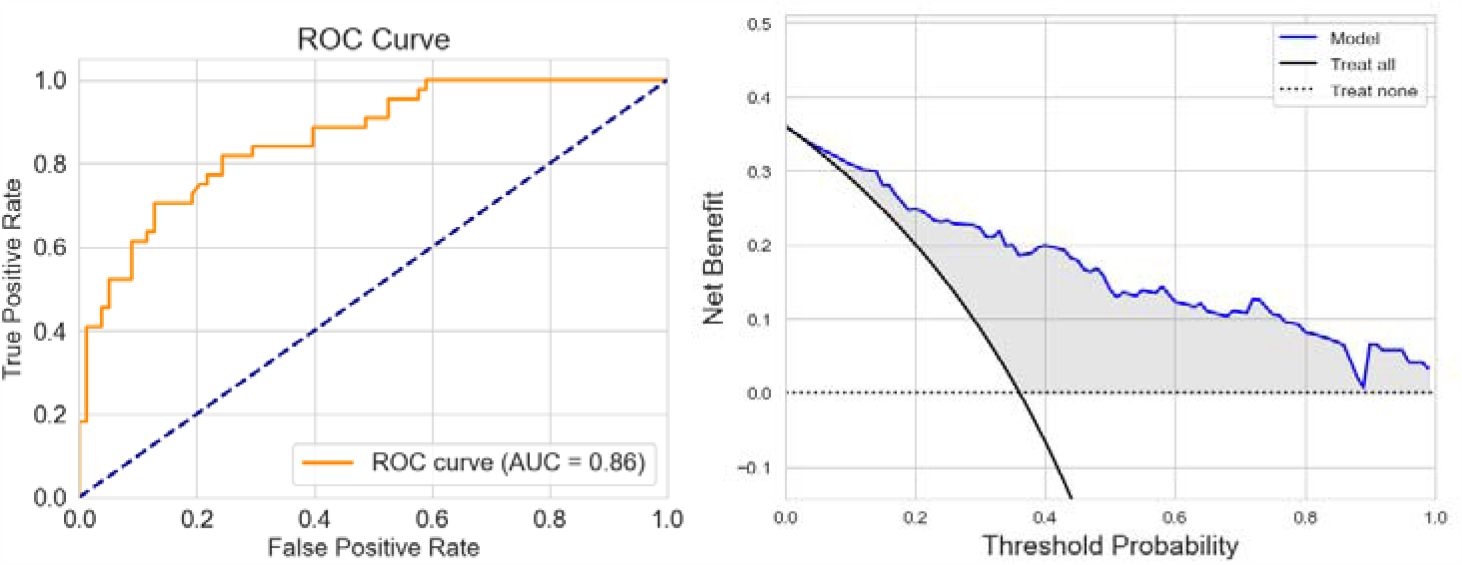
The ROC and DCA curves of the model on the test set.

**Figure 3.**
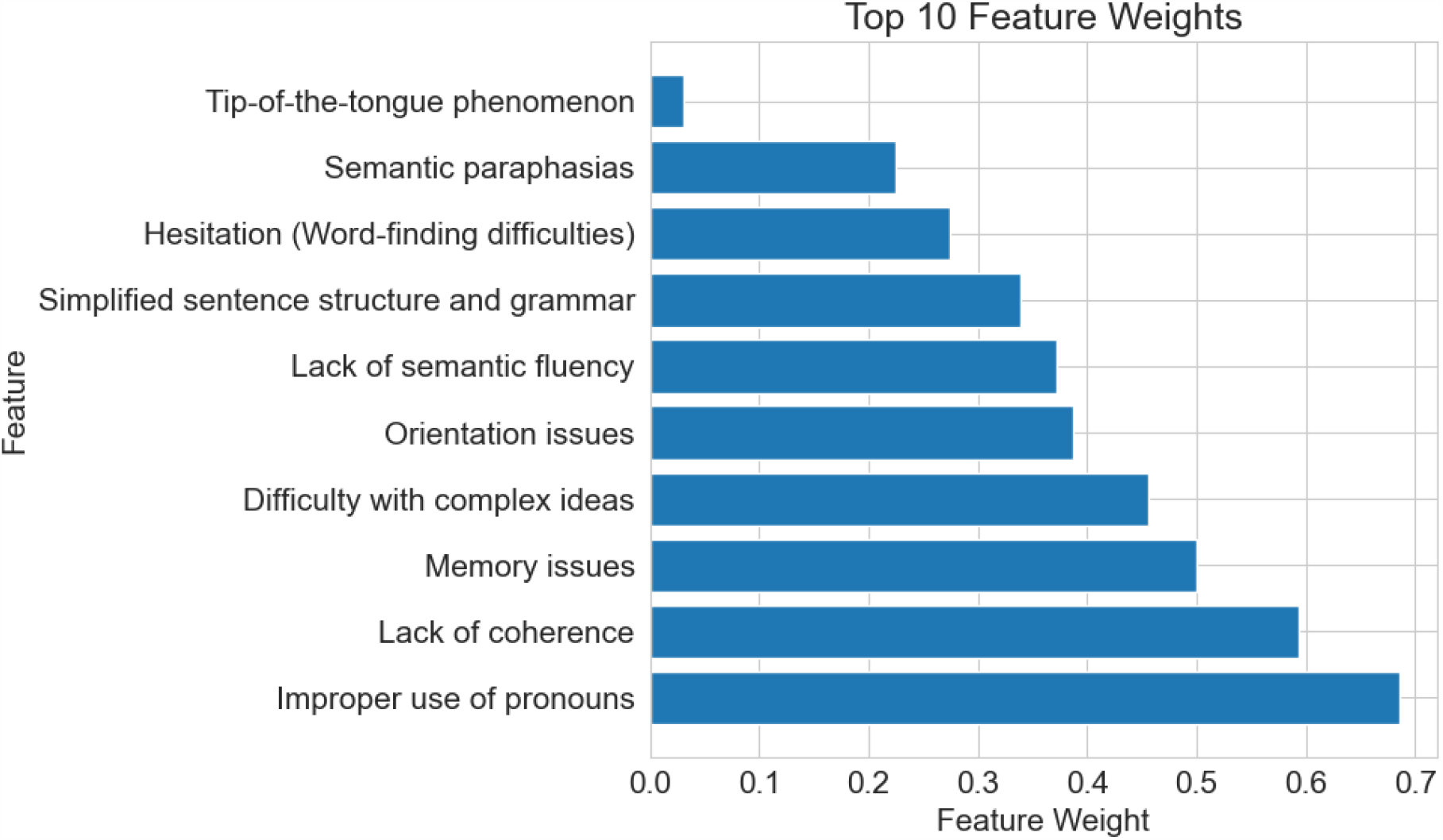
The top ten features contributing the most to distinguishing MCI

### Feature Analysis and Distribution Disparities between MCI and NC

For the top ten ranked features, we used ANOVA to assess whether there were significant differences between patients with mild cognitive impairment (MCI) and those with cognitive normal (NC). This analysis included measures such as median, interquartile range and shape of distribution. The results show that all of these features yielded p-values below 0.05 (see Table 4), indicating the presence of significant differences between them. We also used violin plots (shown in Figure 4) to visually illustrate the distribution of these features between MCI and NC patients. These results help us to understand the differences in the distribution of features between MCI and NC.

**Figure 4.**
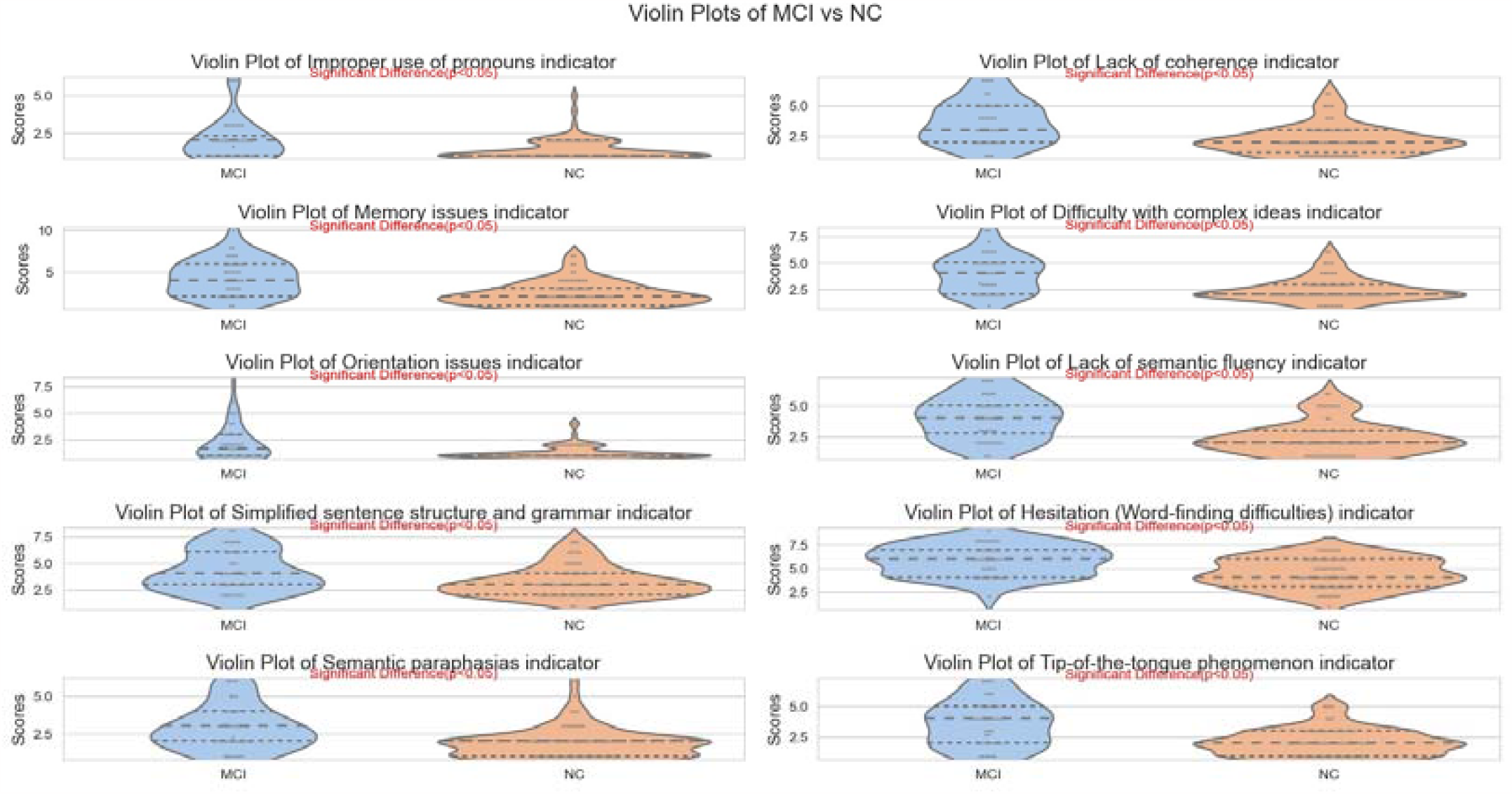
Violin plots for different features.

### Nomogram for Clinical Risk Assessment in MCI Patients

Based on the selection of the top ten weighted features, we constructed a logistic regression model to develop a clinical risk assessment nomogram (Figure 5)[22] for individuals with MCI. A total score of 15 or more indicates potential risk of MCI, with 22.7 being the cut-off (0.43) and a maximum score of 30. These scores help to assess the risk of MCI in patients and provide valuable clinical insight.

**Figure 5.**
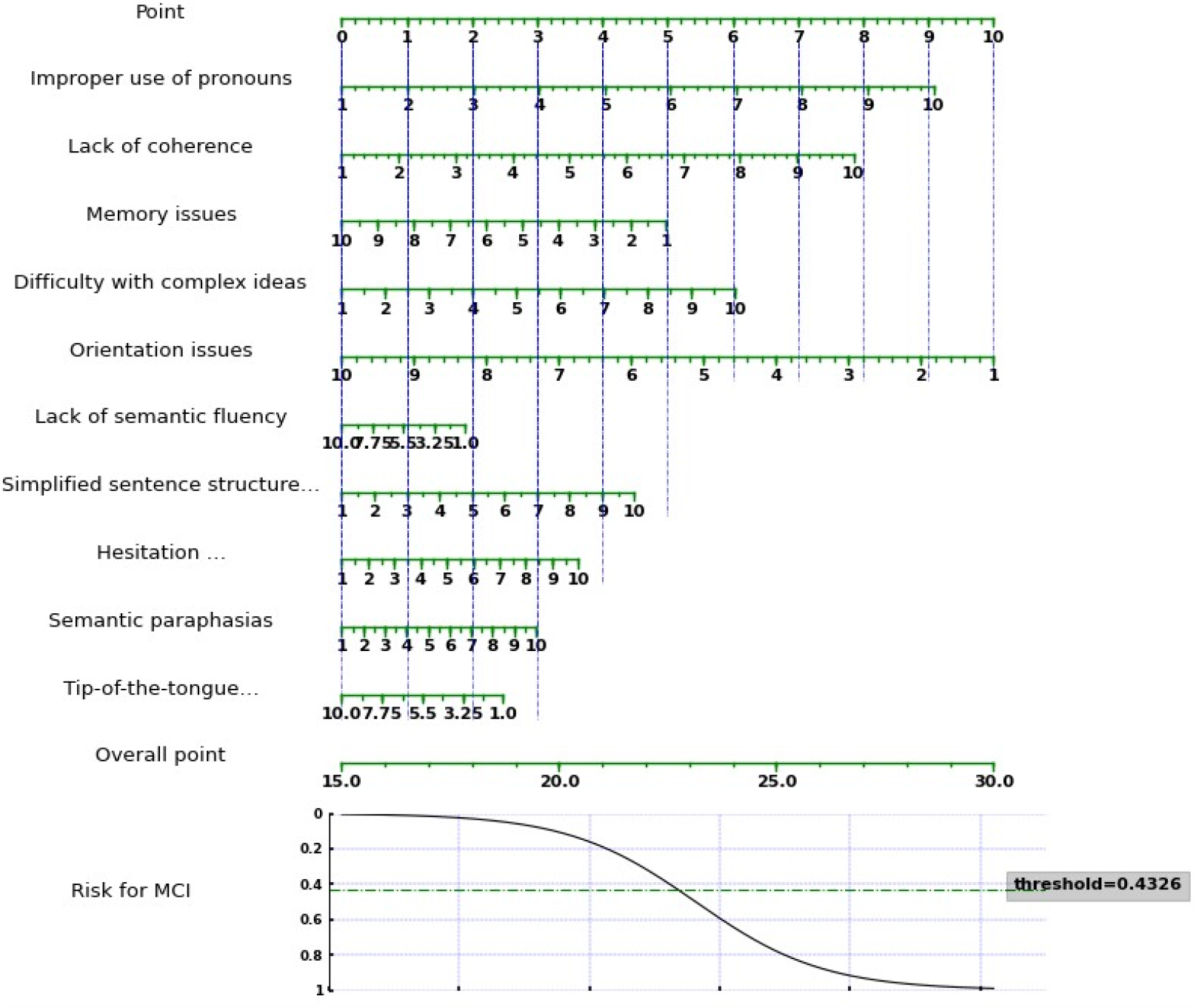
Clinical risk assessment form.

### Psychiatrist evaluation results

In this study we assessed the performance of three neurologists who independently scored four sets of the same corpus. The scoring results showed significant differences between the different neurologists (Table 3). The second neurologist had the highest accuracy of 0.49, closely followed by the third with 0.45 and the first with a relatively low accuracy of 0.41. The difference in scores between neurologists was statistically significant (p<0.01). However, the difference in each neurologist’s score on the four test corpora was not significant (p> 0.05). This suggests that although there were differences in their specific scores, the overall consistency of the scores was relatively good.

**Table 3.** Neurologist’s judgement of MIC.

| model | TP | FP | FN | TN | SEN | SPE | PPV | NPV | PLR | NLR | Accuracy | P(N vs N) | P(G) |
| --- | --- | --- | --- | --- | --- | --- | --- | --- | --- | --- | --- | --- | --- |
| #1(G_1) | 39 | 90 | 27 | 18 | 0.59 | 0.17 | 0.30 | 0.40 | 0.71 | 2.45 | 0.33 |  |  |
| #1(G_2) | 51 | 82 | 15 | 26 | 0.77 | 0.24 | 0.38 | 0.63 | 1.02 | 0.94 | 0.44 | <0.01 | >0.05 |
| #1(G_3) | 49 | 83 | 17 | 25 | 0.74 | 0.23 | 0.37 | 0.60 | 0.97 | 1.11 | 0.43 |  |  |
| #1(G_4) | 49 | 81 | 17 | 27 | 0.742 | 0.25 | 0.376 | 0.61 | 0.989 | 1.03 | 0.436 |  |  |
| #1 | 47.00(5.42) | 84.00(4.08) | 19.00(5.42) | 24.00(4.08) | 0.71(0.08) | 0.22(0.04) | 0.36(0.04) | 0.56(0.11) | 0.92(0.14) | 1.39(0.72) | 0.41(0.05) |  |  |
| #2(G_1) | 34 | 58 | 32 | 50 | 0.52 | 0.46 | 0.37 | 0.61 | 0.96 | 1.05 | 0.48 |  |  |
| #2(G_2) | 34 | 56 | 32 | 52 | 0.52 | 0.48 | 0.38 | 0.62 | 0.99 | 1.01 | 0.49 | <0.01 | >0.05 |
| #2(G_3) | 30 | 61 | 36 | 47 | 0.46 | 0.44 | 0.33 | 0.57 | 0.81 | 1.25 | 0.44 |  |  |
| #2(G_4) | 38 | 54 | 28 | 54 | 0.58 | 0.50 | 0.41 | 0.66 | 1.15 | 0.85 | 0.53 |  |  |
| #2 | 34.00(3.27) | 57.25(2.99) | 32.00(3.27) | 50.75(2.99) | 0.52(0.05) | 0.47(0.03) | 0.37(0.03) | 0.61(0.04) | 0.98(0.14) | 1.04(0.17) | 0.49(0.04) |  |  |
| #3(G_1) | 35 | 75 | 31 | 33 | 0.53 | 0.31 | 0.32 | 0.52 | 0.76 | 1.54 | 0.39 |  |  |
| #3(G_2) | 41 | 67 | 25 | 41 | 0.62 | 0.38 | 0.38 | 0.62 | 1.00 | 1.00 | 0.47 | <0.01 | >0.05 |
| #3(G_3) | 38 | 73 | 28 | 35 | 0.58 | 0.32 | 0.34 | 0.56 | 0.85 | 1.31 | 0.42 |  |  |
| #3(G_4) | 45 | 66 | 21 | 42 | 0.68 | 0.39 | 0.41 | 0.67 | 1.12 | 0.82 | 0.50 |  |  |
| #3 | 39.75(4.27) | 70.25(4.43) | 26.25(4.27) | 37.75(4.43) | 0.6(0.06) | 0.35(0.04) | 0.36(0.04) | 0.59(0.07) | 0.93(0.16) | 1.17(0.32) | 0.45(0.05) |  |  |

**Table 4.**
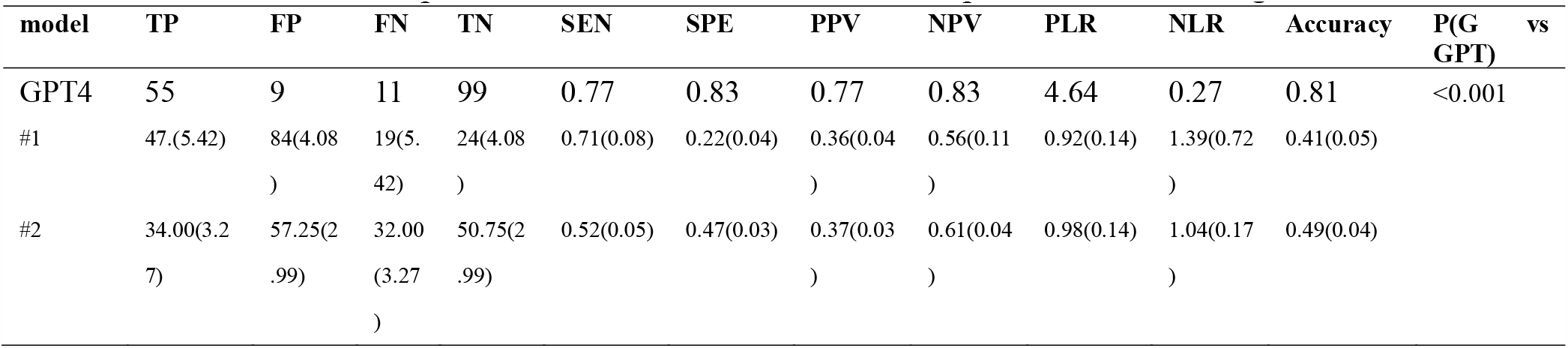
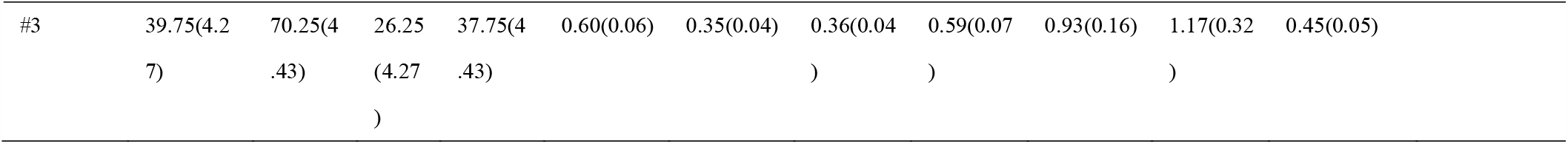
Comparison of results between GPT-4 and professional neurologists.

### Psychiatrists and GPT-4 evaluation

In our study, the diagnostic results of three neurologists were compared with the results of the GPT-4 model, which we had specially trained for this purpose (Table 4). The GPT-4 model achieved an accuracy rate of 0.80, significantly higher than that of the neurologists, which ranged from 0.41 to 0.45. This difference is statistically significant at p < 0.001. These results highlight that the neurologists had significantly higher false positive rates of 84, 57 and 70 respectively, in stark contrast to the GPT-4 model’s false positive rate of 9. This discrepancy highlights a tendency for neurologists to misclassify healthy individuals as having a disease. These findings are further illustrated in Figure 6 by box plots, which provide a visual representation of the data and its variance.

**Figure 6.**
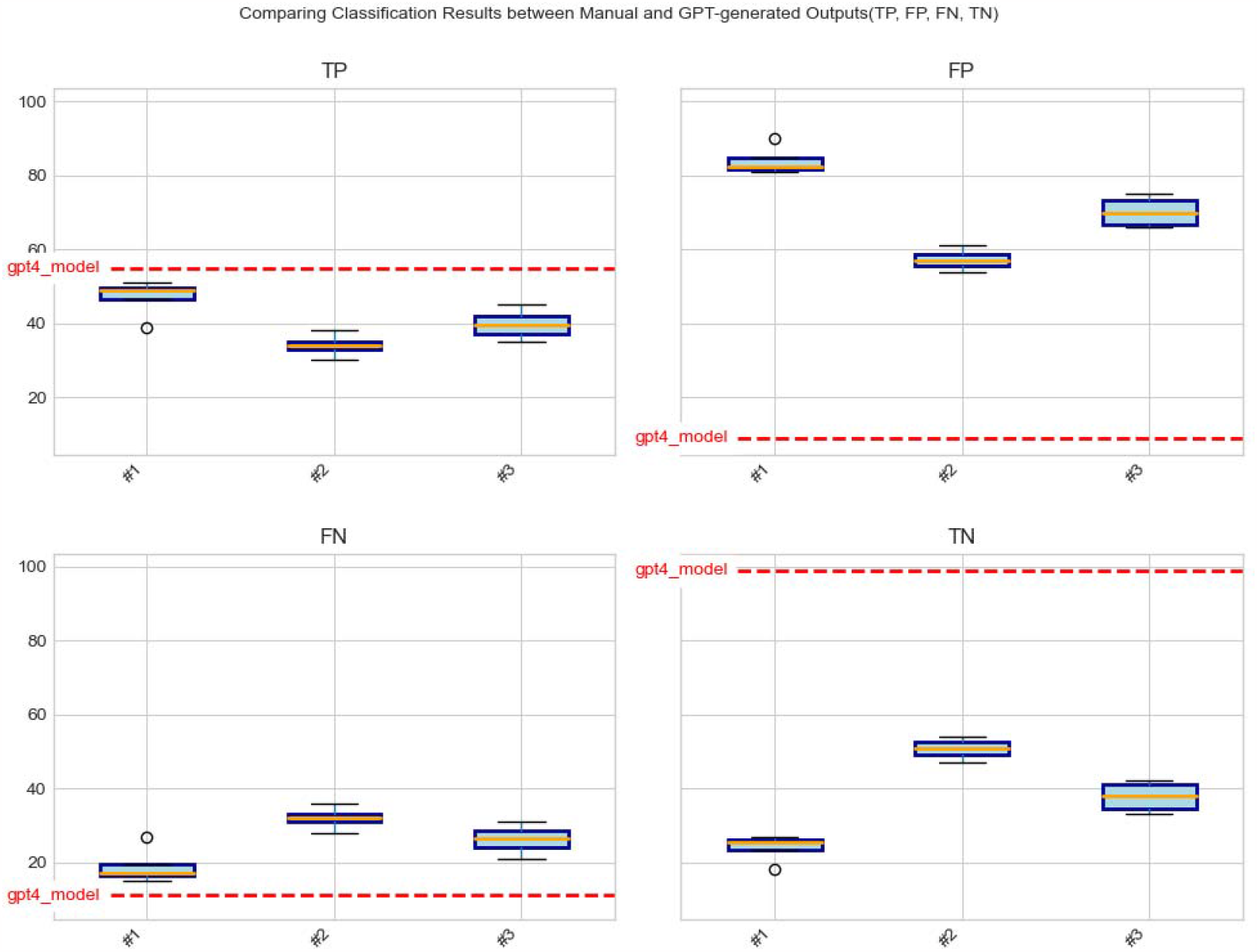
Box plots comparing the results of different neurologists (GPT-4 significantly outperforms human assessments).

## Discussion

Our study reveals that the GPT-4 model emerges as a promising diagnostic aid for MCI, offering an effective and consistent approach for early detection [25]. This capability holds the potential to improve patient outcomes and reduce healthcare expenditures. Since the GPT-4 does not support voice assessment when we trained the GPT-4 model, we only used textual material from MIC patients. In our analysis, the GPT-4 model demonstrated significantly higher accuracy in differentiating between MCI patients and those with NC compared to assessments made by three junior neurologists. Nevertheless, to fully confirm its effectiveness and practicality in real-world MCI diagnosis, further in-depth investigation and rigorous clinical validation of the GPT-4 model are essential.

The purposeful inclusion of junior neurologists in this study mirrors the reality that MCI screenings are often conducted by junior neurologists. This choice provided a valuable opportunity to evaluate the practical utility of the GPT model in a real-world setting and benchmark its performance against human practitioners. Our findings revealed notable variations in the diagnostic scores assigned by the neurologists (p < 0.01), with a discernible positive correlation between their clinical experience and diagnostic accuracy. Conversely, the consistency of neurologists’ assessments for the same patient across various groups did not show a statistically significant difference (p > 0.05), underscoring a high level of rater consistency and reliability.

To enhance our understanding of how the GPT-4 model evaluates MCI and Normal Cognition (CN), we analyzed the feature weights (coef_weight) returned by the model [26]. This analysis identified the top ten features that significantly contribute to differentiating MCI. These linguistic features underscore the importance of specific language elements in detecting MCI, including incorrect pronoun use, lack of coherence, memory problem, difficulty with complex concepts, orientation challenges, diminished semantic fluency, simplified sentence structure and grammar, hesitancy (trouble in finding words), and semantic paraphasia. These findings are in line with previous studies [27,28] and enhance our understanding of the GPT-4 model’s role in MCI assessment. This information will bolster healthcare providers’ confidence in utilizing the model.

Additionally, we developed a clinical risk assessment nomogram based on these ten features. This nomogram serves as a practical tool for clinicians to assess MCI and stratify patient risk levels [29]. It simplifies the process of identifying individuals at high risk for MCI and aids in directing targeted interventions and care strategies. However, this nomogram requires further validation and evaluation. Future research should explore the linguistic features identified in this study more thoroughly, which could uncover linguistic markers indicative of cognitive impairment.

## Limitations

This study has several limitations that need to be considered. Firstly, our analysis was based on publicly available text and speech data. While these data sets are anonymized, it’s crucial to recognize that they might not fully capture the diversity and complexity inherent in real-world clinical scenarios. The limited voice information, which only involved the picture description and/or storytelling, but lack other routine information for clinical diagnosis of dementia, including medical history taking, physical examination, and laboratory tests etc. The incomplete information often results in the inaccurate diagnosis for physicians such as curbside consultations versus formal consultations [30,31]. Future research should aim to include more diverse and comprehensive data sets to improve the model’s applicability across varied contexts. Secondly, the cohort of neurologists involved in this study was relatively small and not native speakers with limited clinical experience. Although the included junior neurologists are proficient in English, and this setup aligns with the practical context of MCI screening, it is important to note that the results may not accurately represent the capabilities of more seasoned practitioners. Future research efforts should involve a larger and more experienced group of neurologists to validate the efficacy of the GPT-4 model in a broader clinical setting [32,33]. Thirdly, our study did not investigate potential biases within the GPT-4 model. It is essential to rigorously evaluate the model’s performance across different populations and to scrutinize it for any inherent biases. Additionally, extending the model’s applicability to other languages and demographic groups is a critical area for future exploration. Finally, due to the unavailability of GPT-4 for speech recognition during the course of our study, our analysis was confined to textual data. Future studies plan to integrate both text and speech data, expanding the scope of recognition and enhancing the model’s utility in clinical assessments.

## Conclusion

Our findings indicate that the GPT-4 model, upon successful training, exhibits potential as a valuable instrument for screening individuals with Mild Cognitive Impairment (MCI). Demonstrating superior accuracy and consistency, it outperforms junior neurologists in preliminary assessments. However, further research and clinical validation are needed to assess the practical applicability of AI models such as GPT-4 in MCI diagnosis.

## Data Availability

The DementiaBank dataset utilized in this study is secured by a password and access is limited to members of the DementiaBank Consortium Group. To obtain permission to access this dataset, one must become a member of the DementiaBank Consortium Group. Informed consent was obtained from all subjects in accordance with the DementiaBank guidelines.

## Acknowledgements

All authors thank DementiaBank for corpus support. The data was provided by DementiaBank and partly supported by NIH AG03705 and AG05133.

## Author Contributions

JL and SL conceived the study. JL, HY, RW, CW, HG, HC, and SL and performed the data analysis, interpreted the results, and drafted the manuscript. All authors revised the manuscript. All authors read and approved the final manuscript.

The article was written entirely by human. All authors are responsible and accountable for the originality, accuracy, and integrity of the work.

